# Transducin: an open-source pipeline recovering SNOMED-CT coded measurements from the undocumented Optopol .OPT and Zeiss Cirrus private-tag formats as DICOM Structured Reports

**DOI:** 10.64898/2026.07.14.26357256

**Authors:** JN Jaurrieta Hinojos, JL Palomares Ordonez, JF Chacon Hinojos, MA Folgueras Batres

## Abstract

**Background:** Quantitative optical coherence tomography (OCT) measurements are essential for retinal disease monitoring, yet leading vendors store acquisition data in undocumented proprietary formats or encode measurements exclusively in private DICOM tags inaccessible to open systems.

**Methods:** We present Transducin, an open-source Python library that reverse-engineers, through systematic binary analysis cross-referenced against DICOM-exported metadata from the same studies, the undocumented Optopol REVO FC130 and REVO 60 .OPT binary format and extracts quantitative measurements from Zeiss Cirrus HD-OCT 500 private DICOM tags, generating TID 1500 Structured Reports with SNOMED-CT coded findings for both platforms. A novel finding is that a scan-geometry parameter recorded by the OCT device (OCTPARAMS tag 23, the horizontal position of the foveal or disc reflex relative to scan center) encodes ocular laterality through its arithmetic sign — an anatomical consequence of the fovea’s nasal position relative to the optic disc — enabling automated laterality inference that requires no operator data entry or filename convention.

**Results:** The validation corpus comprised 475 Optopol .OPT files across two clinical sites — 387 at Site A (Chihuahua; 73 patients) and 88 at Site B (Querétaro) — spanning three device models and four SOCT software versions, with 100% parse success across nine acquisition types. Cross-version compatibility was confirmed across SOCT versions 11.5.0– 21.5.0, spanning approximately eight years of software development. The Zeiss Cirrus pipeline generated TID 1500 SRs for all 41 applicable studies (100%), yielding CMT 203–630 µm and RNFL 53–123 µm across a clinically representative range. The laterality-inference hypothesis was validated across 18 files from two device models and two software versions with 100% accuracy.

**Conclusions:** Transducin provides the first publicly documented specification of the Optopol .OPT format and the first open-source multivendor pipeline generating SNOMED-CT coded DICOM Structured Reports from both Optopol REVO and Zeiss Cirrus devices — closing a gap explicitly confirmed by both manufacturers’ own documentation. The code is available at https://github.com/oftalmos-org/transducin (Apache License 2.0).

## 1. INTRODUCTION

Optical coherence tomography (OCT) has become an indispensable diagnostic tool in ophthalmic practice, enabling non-invasive, high-resolution cross-sectional imaging of retinal microstructure. The Optopol REVO FC130 (Optopol Technology Sp. z o.o., Zawiercie, Poland) is a spectral-domain OCT system widely deployed in clinical settings across the SOCT software platform family, which encompasses nine device models from the SOCT Copernicus REVO to the REVO HR, each storing acquisition data in a proprietary binary format with the .OPT extension. The same .OPT format is used for the multimodal capacities the REVO displays (OCT, OCT angiography, topography, biometry) as well as by other Optopol devices as the perimeters. Despite the clinical ubiquity of these devices, the internal structure of the .OPT format has remained entirely undocumented, an opaque barrier that limits interoperability with open Picture Archiving and Communication System (PACS) infrastructure, hinders research reuse, and creates vendor dependency for clinicians and institutions seeking to integrate OCT data into modern clinical workflows.

The DICOM standard was designed precisely to address this fragmentation, providing a universal framework for medical image exchange and structured clinical reporting. While Optopol publishes a DICOM Conformance Statement for the REVO family (version 21.1.2, July 2025), this document describes only the device’s native export capabilities and explicitly states that no structured coded terminology is used in any export modality (§7.3.5), and that no private DICOM attributes carry additional clinical data (§7.3.4) [1]. No documentation of the .OPT binary format exists in any public registry, including IEEE Xplore, PubMed, arXiv, or SPIE Digital Library; nor is the format supported by any known open-source OCT parsing library, including OCT-Converter [2] or eyepy [3]. The Zeiss Cirrus HD-OCT (Carl Zeiss Meditec, Jena, Germany) represents a distinct but equally consequential interoperability challenge. Unlike the REVO FC130, the Cirrus does export DICOM-compliant images; however, its quantitative measurements, however, are encoded exclusively in private DICOM tags under the vendor-specific coding scheme 99CZM, without SNOMED-CT coded terminology or DICOM Structured Report (SR) generation. Central macular thickness (CMT), Early Treatment Diabetic Retinopathy Study (ETDRS) grids, retinal nerve fiber layer (RNFL) sectors, and optic disc parameters are thus inaccessible to any standards-compliant clinical decision support system or research pipeline. This dual absence of a documented format for Optopol and of machine-readable coded measurements for Cirrus constitutes a concrete clinical informatics gap. Quantitative measurements critical to retinal practice exist within both device families’ data files but are inaccessible to open systems. The gap is particularly consequential in resource-limited clinical settings, where self-hosted open infrastructure represents the only viable path to PACS integration and longitudinal data reuse. This fragmentation is not unique to Optopol and Zeiss: a 2025 systematic review of retinal imaging standardization concluded that no universal, one-to-one converter exists for transforming non-compliant ophthalmic imaging formats into DICOM, and that converters must instead be tailored to specific imaging protocols and devices [4] — precisely the device-specific approach adopted here for both platforms.

These two device families were selected for study because they represent the imaging equipment available to the study site: a REVO FC130 for day-to-day retinal imaging, and a Cirrus HD-OCT retained for cross-reference and continuity of care. In practice, moving a patient’s OCT record between the two devices meant their studies could not be viewed within a single PACS — each vendor’s proprietary export format required its own dedicated viewer, fragmenting a single patient’s longitudinal imaging record across incompatible silos. This first-hand experience of vendor lock-in, rather than a top-down survey of the OCT device market, motivated the reverse-engineering effort described here. The legal and ethical basis for this reverse-engineering effort, including licensing terms and vendor notification, is addressed in Section 4.8.

We present Transducin, an open-source Python library that addresses both gaps: for the first time, it documents the internal structure of the Optopol .OPT binary format, implements a complete parser for its clinical content, and additionally implements a Zeiss Cirrus pipeline that extracts quantitative measurements from private DICOM tags and generates TID 1500 Structured Reports (SRs) — DICOM’s standard template for coded, machine-readable quantitative measurement reports — with SNOMED-CT coded findings. The remainder of this paper is organized as follows: Section 2 describes the .OPT format specification, the Transducin conversion pipelines, and validation protocols. Section 3 presents validation results. Section 4 discusses implications for ophthalmic data interoperability and future directions.

This work is a software development and validation study. All validation data are fully de-identified and no patient data are distributed. The broader ophthalmology software ecosystem of which Transducin is the first published module operates under the IEEE/UL 2933-2024 TIPPSS framework [5] for clinical IoT security. An institutional ethics committee (Hospital Angeles Chihuahua) waived ethical approval for this work. This manuscript treats the standard DICOM image generation, the DICOM SR encoding, and the PACS-integrated delivery pipeline as a single, necessarily coupled contribution: format decoding has no clinical utility in isolation, and its correctness is demonstrated here only by end-to-end delivery of coded measurements into a standards-compliant PACS, which is the validation performed throughout Section 3.

## 2. METHODS

### 2.1. OPT Format Analysis

Development of Transducin began against .OPT files generated by SOCT software version 11.5.x, provided by a collaborating retinal practice in Querétaro, México (Site B, Section 2.8), prior to the primary author’s acquisition of a REVO FC130 unit. This cross-version origin proved consequential: the chunk-container architecture and segmentation layer identifiers were subsequently confirmed identical across SOCT versions 11.5.0, 11.5.3, and 21.1.2, spanning approximately eight years of software development. The robustness to version variation embedded in the parser’s design reflects this developmental history rather than post-hoc compatibility patching.

Primary binary analysis was conducted on .OPT files generated by a single REVO FC130 unit running SOCT software version 21.1.2 at Site A (Hospital Angeles Chihuahua). The manufacturer-provided DICOM Conformance Statement (v21.1.2, July 2025) [1] and CMDL Interface documentation (revision B, July 2025) [6] were used as reference materials to validate identifier structures and attribute mappings. The .OPT format itself has no published specification; its internal structure was determined through systematic binary analysis, with chunk semantics inferred from cross-referencing extracted values against DICOM-exported metadata from the same studies. Concretely, this involved hex-level inspection of raw .OPT bytes to locate repeating four-character chunk-code markers; iterative decompression attempts (zlib) on candidate chunk payloads to confirm chunk boundaries and compression flags; comparison of decoded numeric fields (e.g., axial length, scan center coordinates) against the corresponding values exported by the same device in companion DICOM files and in the SOCT native software display, to assign semantic meaning to otherwise opaque byte offsets; and, for the one chunk that resisted this process (the patient-identity block), the entropy and XOR-sweep analysis described above (Section 2.1). Binary analysis and software development were carried out by a team combining clinical and technical expertise: two authors are practicing retina specialists with direct clinical familiarity with the acquisition protocols and output data being reverse-engineered, and two are systems engineers, one holding a postgraduate degree in image processing and the other in software engineering (see Author Contributions).

In total, the validation corpus spans four SOCT software versions (11.5.0, 11.5.3, 21.1.2, 21.5.0), three physical Optopol devices (two REVO FC130 units and one REVO 60 unit), and nine acquisition types; full per-type and per-version breakdowns are reported in Tables 3, 4, and 5.

File identification. Every .OPT file begins with a four-byte magic signature: 0×A5 0×A5 0×A5 0×FF. This signature was checked against the GCK File Signature Table (filesig.search.org, the actively maintained successor to Gary Kessler’s File Signatures Table), a public registry of several hundred known file-format magic numbers, with no match found; a search of every file format in existence remains impractical to perform or claim. Files range from approximately 50 MB to 800 MB depending on acquisition type and number of averaging frames. Figure 1 details the architecture of the chunks.

**Figure 1.**
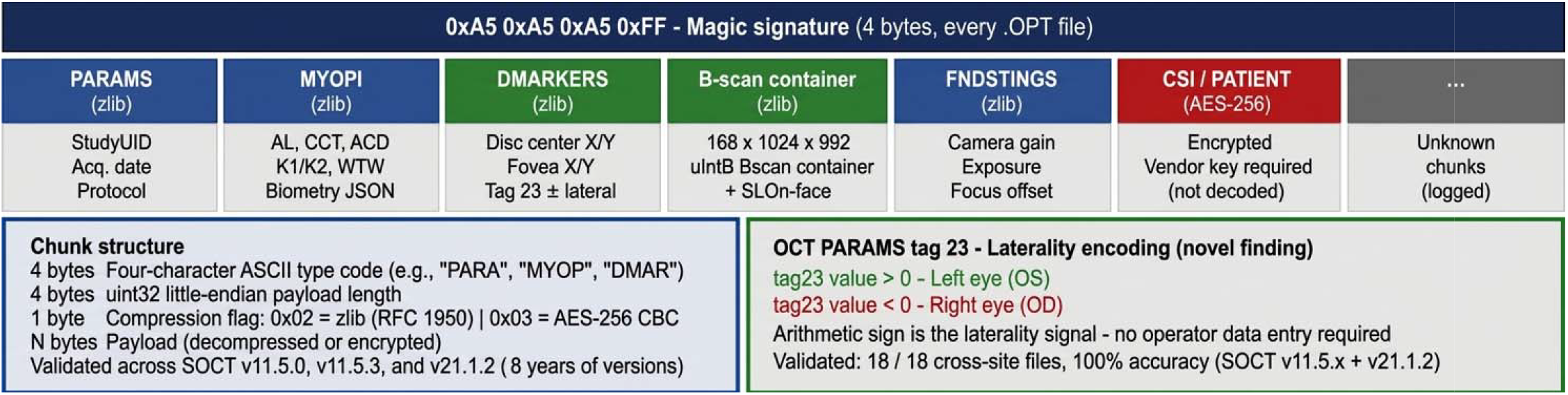
The Optopol .OPT binary format. Each file begins with a four-byte magic signature (0×A5A5A5FF) followed by a sequence of named chunks. Clinical data chunks use zlib compression (0×02); the patient identity block uses a strong symmetric encryption of unconfirmed cipher and mode, with simple single-byte XOR ruled out (0×03). Novel finding: OCTPARAMS tag 23 encodes ocular laterality through its arithmetic sign, validated against 18 cross-site files with 100% accuracy.

Container architecture. Following the magic bytes, the .OPT file is organized as a sequence of named chunks. Each chunk begins with a four-character ASCII code (the chunk type identifier) followed by a 32-bit little-endian integer specifying the payload length, and then the payload itself. This chunk-based architecture is consistent across all scan types and software versions tested.

Compression modes. Two compression modes were identified, distinguished by a single byte flag at a fixed offset within each chunk header: 0×02 indicates zlib (RFC 1950) compression, and 0×03 indicates strong symmetric encryption of unconfirmed cipher and mode. Clinical data chunks use zlib compression and are fully decodable without proprietary keys. The patient identity block uses the encrypted format and cannot be decoded without a vendor-specific key. Its cipher and mode were not identified. Shannon entropy of the encrypted block measured 7.988 bits/byte, consistent with strong encryption rather than a simple substitution scheme, and an exhaustive sweep of all 256 single-byte XOR keys ruled out simple XOR obfuscation. No attempt was made to break the encryption. Patient demographics are instead sourced from companion DICOM files or from the electronic health record (EHR) via the NOEL ID protocol (Section 2.6).

### 2.2. Chunk Specification

In SOCT versions prior to 21.x, biometry data are stored under a distinct chunk schema (WTW, TPR, S0–S9) rather than the MYOPI JSON structure; this variation is documented as a known limitation (Section 4.3). Chunks with unrecognized four-character codes are preserved in a passthrough buffer and logged with their offset and length in a JSON manifest. Table 1 summarizes the chunks identified in this work, with their four-character codes, compression modes, payload formats, and clinical content.

### 2.3 Laterality Inference and Scan Type Discrimination: Methodology

Laterality from OCTPARAMS tag 23. The OCTPARAMS binary structure includes a floating-point value at a fixed offset (tag index 23) representing the horizontal position of the foveal reflex or disc center relative to the scan center, in millimeters. To test whether this value encodes laterality, it was cross-referenced against 13 files with independently ground-truthed laterality labels (from REVO 60 files with explicit _L_/_R_ filename tokens — an initial single-device test set; full validation across both device models totals 18 files and is reported in Section 3.6), on the anatomical hypothesis that a sign-based relationship should exist: the fovea is located nasal to the disc in both eyes, so in a standard scanning laser ophthalmoscope (SLO) image the nasal direction is to the right for the right eye (negative horizontal offset) and to the left for the left eye (positive offset). This parameter is computed by the REVO firmware from the SLO acquisition geometry and, if the hypothesis held, would provide a laterality signal independent of operator data entry. Validation of this hypothesis across scan types, device models, and software versions is reported in Section 3.6.

Scan type discrimination via chunk presence. Rather than relying solely on filename tokens or OCTPARAMS-derived parameters, Transducin implements a deterministic hierarchy for scan type classification based on which chunks are present in the .OPT file:

1. ANGPRV chunk present → acquisition type = angio (OCTA flow acquisition)
2. DMARKERS chunk present → acquisition type = optic_nerve (optic disc acquisition). The DMARKERS chunk encodes disc center coordinates and is present exclusively in disc-centered acquisitions; its absence in macular scans was confirmed across all three device models and all four SOCT versions tested.
3. EYE chunk present without DMARKERS → macular (n_frames ≥ 100) or hd_line (18 frames × 1,024 A-scans, single high-definition line raster)
4. FNDSRECO present without EYE → wide_field or ultra_wide
5. Keyword fallback → filename stem is searched for recognizable protocol keywords (e.g., “biometr”, “macu”, “topo”, “nerv”) when all chunk-based rules fail

### 2.4 B-scan Extraction

The B-scan container payload, after zlib decompression, yields a raw uint8 array of dimensions [168, 992, 1024] for the REVO FC130, reordered to the conventional DICOM frame layout [168, 1024, 992]. The REVO 60 uses a distinct axial depth of 944 pixels, confirming hardware-level differences within the device family that the parser resolves transparently.

The en-face (SLO) image is extracted from the EYE chunk and requires a reshape operation to 1,024 × 1,024 pixels, distinct from the volumetric frames. Acquisition date and time are extracted from the StudyInstanceUID embedded in the PARAMS chunk (format: 1.YYYYMMDDHHMMSS), ensuring that all eight DICOM date/time tags (StudyDate, StudyTime, SeriesDate, SeriesTime, AcquisitionDate, AcquisitionTime, ContentDate, ContentTime) reflect the actual acquisition timestamp rather than the conversion timestamp.

### 2.5 DICOM Object Generation

For each successfully parsed OPT file, Transducin generates DICOM objects according to available chunks: an OphthalmicTomographyImageStorage instance (SOP Class UID 1.2.840.10008.5.1.4.1.1.77.1.5.4) encoding the B-scan volume, and an OphthalmicPhotography8BitImageStorage instance (SOP Class UID 1.2.840.10008.5.1.4.1.1.77.1.5.1) encoding the SLO en-face image when available.

All generated objects include:

⍰ ProtocolName (0018,1030): populated with the internal scan type identifier
⍰ BodyPartExamined (0018,0015): “EYE ANTERIOR SEGMENT” for anterior segment acquisitions, “EYE” for all posterior segment acquisitions
⍰ PixelSpacing (0028,0030): populated in millimeters from the axial and lateral resolutions extracted from the OCTPARAMS chunk, enabling calibrated measurements in downstream viewers
⍰ AnatomicRegionSequence (0008,2218): populated with SNOMED-CT SRT codes — T-AA700 (“Anterior segment of eye”) for anterior segment acquisitions, T-AA610 (“Retina”) for macular and OCTA acquisitions, and T-AA630 (“Optic nerve head”) for disc acquisitions
⍰ SeriesDescription (0008,103E): formatted as “REVO FC130 {ScanType} {Laterality}” (e.g., “REVO FC130 Macular OD”), derived from the classified scan type and the OCTPARAMS tag 23 laterality
⍰ Laterality (0020,0060) and ImageLaterality (0020,0062): populated with “R” or “L” from the same laterality inference

### 2.6 DICOM SR Generation via TID 1500

The DICOM Structured Report is generated using the highdicom library [7] following TID 1500 (Measurement Report). The SR encodes quantitative findings extracted from the .OPT file and computed from the B-scan data, each expressed as a NUM content item with a SNOMED-CT concept code, a Unified Code for Units of Measure (UCUM) unit, and a floating-point value. Table 2 lists the coded findings included in the SR for each acquisition type. For readers unfamiliar with the DICOM SR structure, Figure 2 illustrates the resulting content hierarchy both conceptually (Figure 2A) and for a real macular study (Figure 2B), together with the downstream queryability that this coded representation enables (Figure 2C).

**Table 1.** Identified .OPT chunk types and their clinical content.

| Chunk Code | Compression | Payload Format | Clinical Content |
| --- | --- | --- | --- |
| PARAMS | zlib | XML-like key-value | StudyInstanceUID, acquisition date/time, scan protocol, device serial number |
| MYOPI | zlib | JSON | Axial length, anterior chamber depth, lens thickness, central corneal thickness, K1/K2, pupil diameter, white-to-white |
| OCTPARAMS | zlib | Binary struct | Acquisition geometry including scan center coordinates, FOV, resolution; tag 23 = foveal horizontal position (mm) |
| DMARKERS | zlib | Binary struct | Disc center X/Y coordinates (pixels), fovea center X/Y coordinates |
| FNDSTNGS | zlib | Binary struct | Fundus camera gain, exposure, focus offset, field of view |
| B-scan container | zlib | Raw uint8 array | Volumetric B-scan data, 168 frames × 1,024 A-scans × 992 pixels (FC130); variant dimensions in other models |
| ANGPRV | zlib | Binary | OCTA angiography preview image (presence indicates OCTA acquisition type) |
| FNDSRECO | zlib | Binary | Fundus reconstruction image (wide-field acquisitions) |
| EYE | zlib | Binary | SLO en-face fundus image (all posterior segment acquisitions) |
| EXAM_AI_RE<br>S | zlib | JSON (variable) | Reserved for AI analysis results (vendor-specific, not decoded) |
| CSI | 0x03 (encrypted) | Strong symmetric encryption (cipher/mode unconfirmed) | Vendor cryptographic signature; not decoded |

*†Codes used within TID 1500 Measurement Report, which does not restrict coding scheme. Migration to DICOM Supplement 247 templates (TID 2123/2124) will require Logical Observation Identifiers Names and Codes (LOINC) codes (CID 4285) for macular measurements and DCM codes (CID 4283) for RNFL measurements.*

**Table 2.** SNOMED-CT coded findings in Transducin SR output by scan type.

| Finding | SNOMED-CT Code | UCUM Unit | Source |
| --- | --- | --- | --- |
| Central macular thickness | 422453003 | $\mu\text{m}$ | Segmentation ILM–BM layer boundary |
| ETDRS inner ring superior | 422991002 | $\mu\text{m}$ | 3 mm annulus, superior sector |
| ETDRS inner ring nasal | 422659000 | $\mu\text{m}$ | 3 mm annulus, nasal sector |
| ETDRS inner ring inferior | 422752009 | $\mu\text{m}$ | 3 mm annulus, inferior sector |
| ETDRS inner ring temporal | 422756007 | $\mu\text{m}$ | 3 mm annulus, temporal sector |
| ETDRS outer ring (×4 sectors) | (respective codes) | $\mu\text{m}$ | 6 mm annulus |
| Peripapillary RNFL mean | 422995006 | $\mu\text{m}$ | Circular 3.46 mm scan |
| RNFL | 422995006 | $\mu\text{m}$ | Sector arcs |
| superior/inferior/nasal/temporal |  |  |  |
| mGCIPL minimum | 416798008 | μm | 6-sector minimum ganglion cell |
| Axial length | 400962006 | mm | MYOPI chunk |
| Central corneal thickness | 422183001 | μm | MYOPI chunk |
| Anterior chamber depth | 400956009 | mm | MYOPI chunk |
| Cup-to-disc ratio | 363932005 | {ratio} | DMARKERS + segmentation |
| Image quality index | — | {ratio} | SQI from PARAMS chunk |

**Figure 2.**
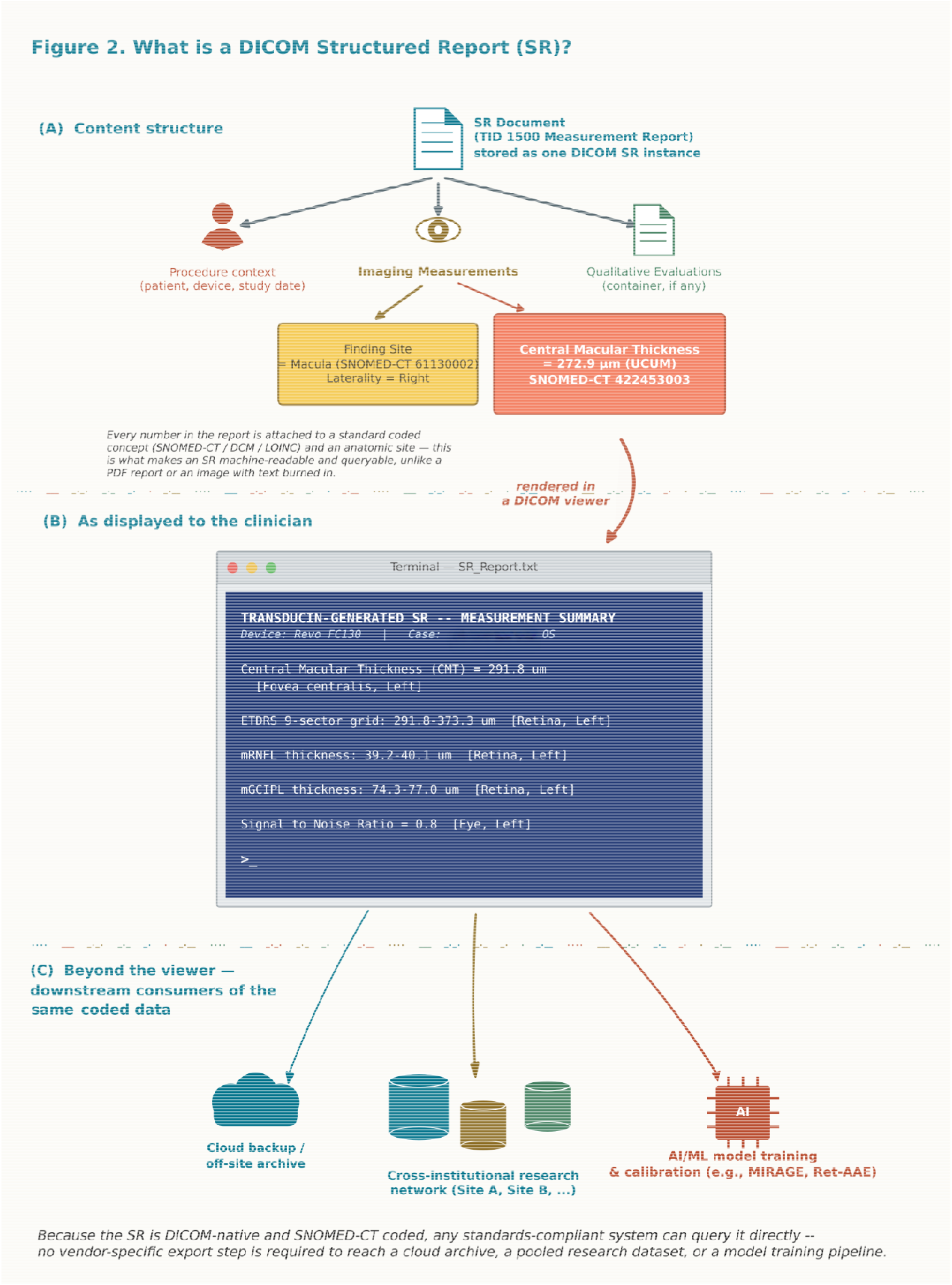
DICOM Structured Report (SR) content structure, from model to downstream use. (A) TID 1500 hierarchy: each finding carries a coded concept (SNOMED-CT/DCM/LOINC), Finding Site, and laterality — making the SR machine-readable and queryable, unlike a PDF or an image with text burned in. (B) Measurement summary from a real Transducin-generated, showing CMT, ETDRS, RNFL, and macular ganglion cell–inner plexiform layer (mGCIPL) sectors collapsed to value ranges for readability. (C) Because the SR is coded and DICOM-native, it is directly queryable beyond the viewer by cloud archives, cross-institutional research datasets, or AI/ML training pipelines, with no vendor-specific export step.

CMT is computed as the mean internal limiting membrane (ILM)–Bruch’s membrane (BM) distance within a 1 mm diameter circle centered on the fovea (radius < 500 µm), expressed in micrometers — equivalent to the ETDRS central subfield (ETDRS-C) as defined in the SOCT native display. Scan Quality Index (SQI) values below 0.4 trigger an automatic warning flag in the SR ContentItems. The provisional 99OFTALMOS private coding scheme is used for findings without a current SNOMED-CT code, registered as a CodingSchemeIdentificationItem per DICOM PS3.3 C.12.1, tagged for replacement upon merging of the planned highdicom pull request (#407).

### 2.7 NOEL ID Protocol

Patient identity propagation is managed via the NOEL ID (Name-Origin Encoded Locally Identification) protocol: a deterministic pseudonym computed locally from a fixed combination of truncated name components and date of birth, never transmitted off-site in reversible form. The exact character-selection and concatenation scheme is intentionally omitted from this manuscript to prevent reconstruction of patient identifiers from publicly available name/birthdate pairs. Because the algorithm is deterministic and applied identically at every site, two devices or institutions computing the identifier for the same patient from the same name and date of birth arrive at the same pseudonym independently, without exchanging identifiable data or relying on a shared national patient identifier; this is what preserves cross-site interoperability (Section 2.8) despite the identifier itself carrying no reversible identifying information. Mexico’s national population registry identifier (CURP, Clave Única de Registro de Población) addresses a related identification problem but is documented to produce inconsistent results in edge cases such as compound surnames, homonyms, or late civil-registry corrections; NOEL ID was designed as a locally-computable, institution-specific identifier for this work rather than as a general replacement for CURP in contexts where it applies.

The resulting identifier travels as DICOM PatientID (0010,0020) through all C-STORE operations. When no NOEL ID is available from the EHR, the parser attempts extraction from the .OPT filename using the regex ^([A-Z]{4})(\d{8})_; if absent, PatientID is left empty rather than populated with a sentinel value such as “UNKNOWN”, to preserve PHI separation from downstream systems.

### 2.8 Deployment Architecture — Optopol REVO FC130

Transducin is deployed on the HP Z2 G1i workstation running Windows 11 hosting the REVO FC130 acquisition software. Two Windows services managed by NSSM run continuously:

- TransducinRevoWatcher monitors the SOCT data directory via filesystem events. On detection of a new .OPT file, it polls for the presence of segmentation layer chunks (BM and TOP) with a configurable timeout (default 180 seconds), addressing a two-stage export behavior in which the REVO FC130 initially writes an incomplete file (∼126 MB) before completing the segmentation pass (∼147 MB). Once segmentation chunks are confirmed present, the pipeline executes: parse → generate DICOM objects → generate SR → C-STORE to Orthanc PACS.
- TransducinPTSWatcher polls the Orthanc REST API every 5 minutes for new OphthalmicVisualField studies without an associated SR, then invokes pts925_extractor.py to generate TID 1500 visual field SRs from the embedded DICOM OPV data.

### 2.9 Cross-Version Validation Protocol

.OPT files were solicited from a collaborating retinal practice in Querétaro, México (Site B), operating two Optopol devices under SOCT software versions predating the primary corpus by approximately eight years: a REVO 60 running SOCT 11.5.3 and a REVO FC130 running SOCT 11.5.0. Site B files were processed using the identical parser and validation script as the primary corpus, with no parser modifications made prior to the cross-version run. De-identified results are archived at validation/site_b_results_anon.csv in the public repository. Quantitative measurement accuracy was assessed by comparing Transducin output against values displayed in the SOCT native software: CMT was compared for two representative macular scans from the primary corpus (same patient, OD and OS); RNFL sector values were compared for 10 studies (94 sector values total); and axial length was compared for 39 biometry files.

### 2.10 Zeiss Cirrus HD-OCT Pipeline

The Zeiss Cirrus HD-OCT 500 exports DICOM-compliant studies but stores quantitative measurements exclusively in private tags under the vendor coding scheme 99CZM (Carl Zeiss Meditec). The pipeline identifies analysis files via the SpatialRegistration SOP Class (1.2.840.10008.5.1.4.1.1.66), which the Cirrus co-exports alongside each volumetric study and which carries the quantitative analysis results in private tags (0073,1150) for ILM layer boundaries and (0073,1255) for ONH XML data.

The acquisition type is identified from PerformedProtocolCodeSequence (0040,0260) item 0, CodeValue field, using the Zeiss private coding scheme: SD-MTA (Macular Cube 512×128) and SD-GOUA (Optic Disc Cube 200×200) are the two acquisition types for which quantitative analysis data are present. HD 5 Line Raster (SD-S51) and anterior segment scans generate DICOM image objects but no analysis files and therefore produce no SR output — this is a Cirrus platform constraint, not a Transducin limitation. Notably, PerformedProtocolCodeSequence[0].CodeMeaning may contain appended null bytes in approximately 3% of files; CodeValue is used as the primary key to avoid this artifact.

Layer boundary arrays are decoded as uint16 little-endian, with dimensions inferred dynamically from payload size: 128×512 pixels for macular acquisitions (n_pixels = 65,536) and 200×200 pixels for disc acquisitions (n_pixels = 40,000). This dynamic inference resolved a silent parsing failure that affected all disc studies when a fixed macular shape was assumed — a format variation not documented in the Zeiss DICOM Conformance Statement. CMT and ETDRS grids are computed from the macular ILM/BM arrays; RNFL sectors and disc morphometry are extracted from the ONH XML embedded in the disc analysis file.

Cirrus pixel data (JPEG 2000-encoded B-scan and fundus frames) is stored in an obfuscated form undocumented in the Zeiss DICOM Conformance Statement: every seventh byte is XORed with a fixed key, and the 253-byte JP2 codestream header is relocated to approximately three-fifths of the frame length. The reversal procedure applied here was adapted, not independently derived, from a public technical discussion on the pydicom project’s GitHub repository [8]; frames are subsequently transposed post-decoding so that depth runs along Rows per OCT convention. This pixel-data reversal step is distinct from, and a precondition for, the private-tag semantic extraction described above, which constitutes this work’s original contribution to the Cirrus pipeline.

The generated TID 1500 SR uses the same SNOMED-CT codes as the Optopol pipeline (Table 2), enabling consistent downstream query and retrieval regardless of OCT vendor.

## 3. RESULTS

### 3.1 Primary Validation Corpus — Optopol REVO FC130

The primary validation corpus comprised 387 .OPT files representing 73 patients acquired at a single retinal specialty practice (Site A, Hospital Angeles Chihuahua) between February and April 2026 under SOCT software versions 21.1.2 and 21.5.0. Files spanned all supported acquisition types across both eyes (Table 3). Patients’ age ranged from 18 to 84 years (median 61). The OCT sample represented included age-related macular degeneration (wet and dry), diabetic macular edema, diabetic retinopathy, glaucoma, epiretinal membrane, vitreomacular traction, and healthy controls. Figure 3 shows a native SOCT display from REVO FC130 against the validation of Transducin output files in OHIF. The scan-type classification hierarchy (Section 2.3) correctly classified all 475 files in the combined Site A + Site B corpus, including files with non-standard filename conventions such as “oct macular.opt” and “oct nervio optico.opt”.

**Figure 3.**
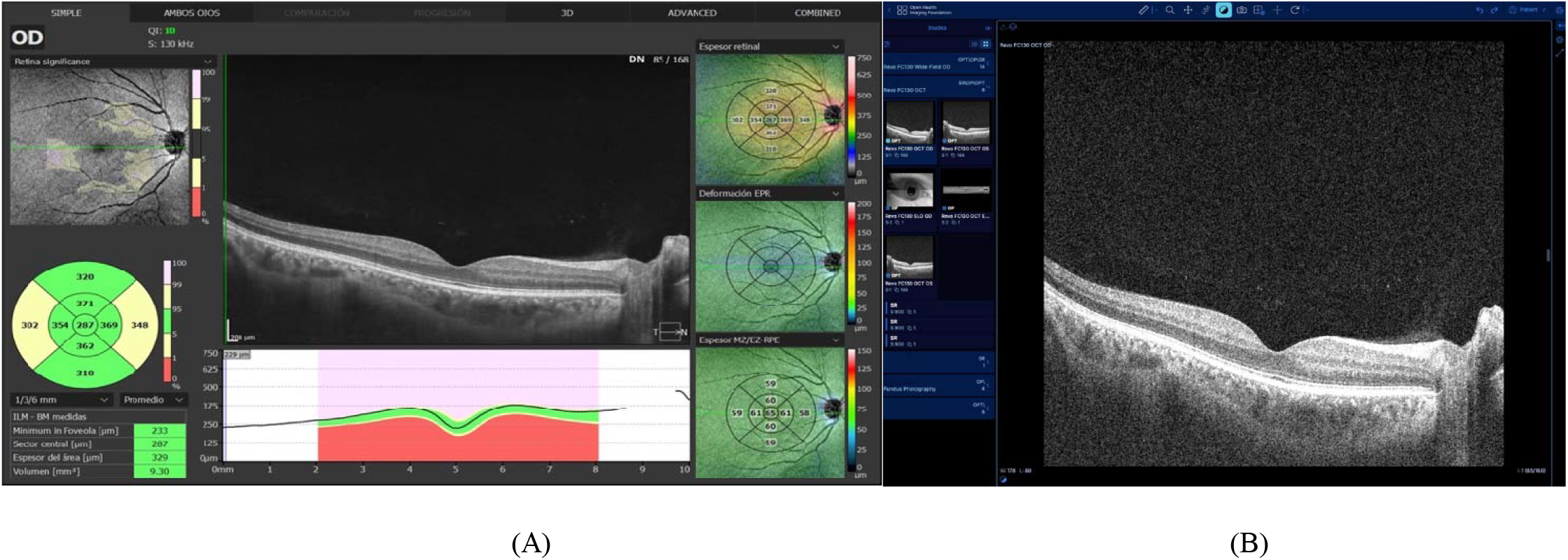
Validation of Transducin output against native SOCT display. (A) SOCT software native display of macular cube showing the ETDRS 9-sector macular thickness grid (Sector central = 287 µm), B-scan with ILM–BM segmentation boundaries, and Retina Significance map. (B) OHIF Viewer rendering the same study after Transducin conversion, showing the OphthalmicTomographyImageStorage volume (168 B-scans, frame 85/168), SLO en-face image, and associated Structured Reports (SR ×3) retrieved from Orthanc PACS.

**Table 3.**
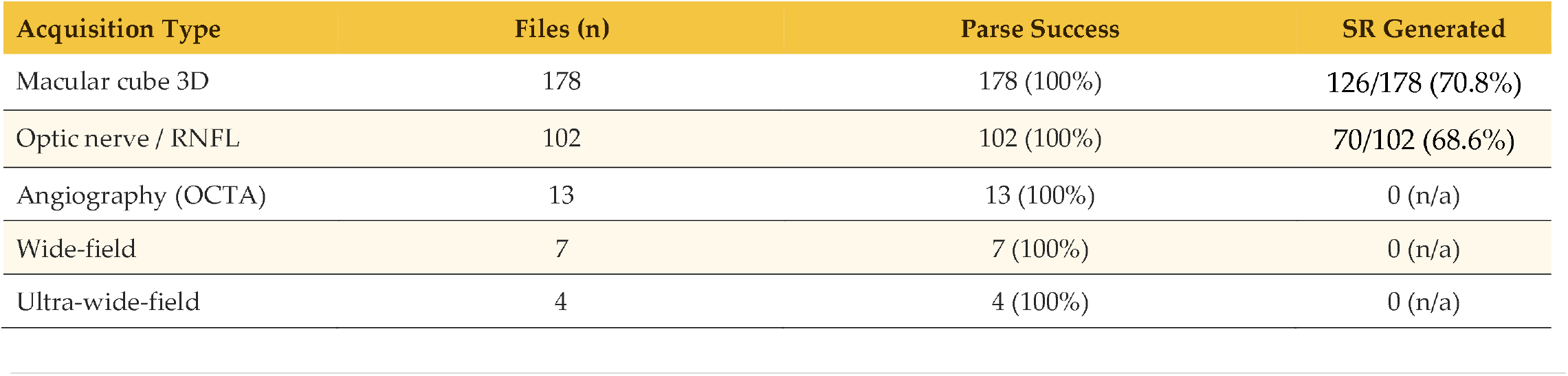
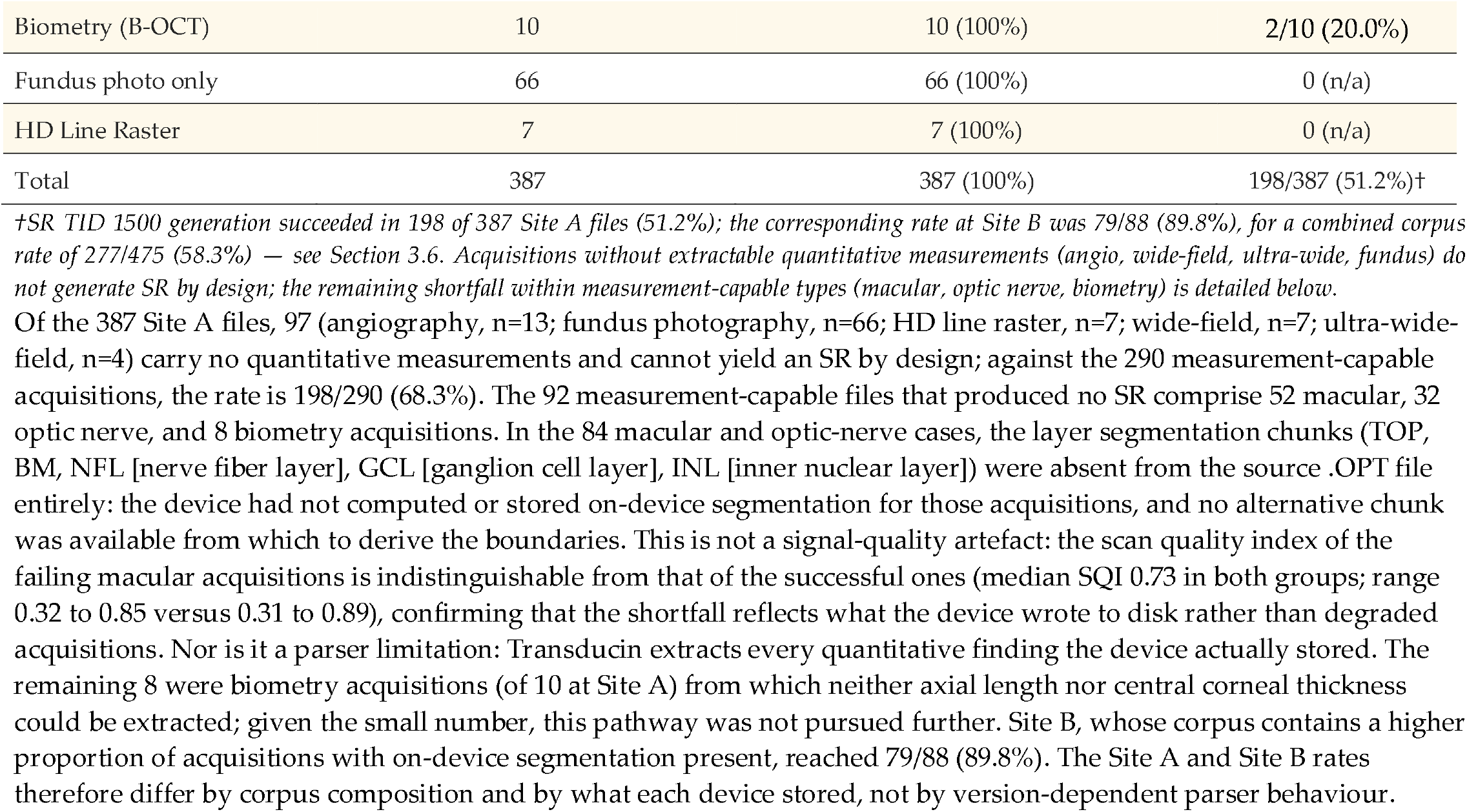
Primary validation corpus composition by acquisition type.

Of the 387 Site A files, 97 (angiography, n=13; fundus photography, n=66; HD line raster, n=7; wide-field, n=7; ultra-wide-field, n=4) carry no quantitative measurements and cannot yield an SR by design; against the 290 measurement-capable acquisitions, the rate is 198/290 (68.3%). The 92 measurement-capable files that produced no SR comprise 52 macular, 32 optic nerve, and 8 biometry acquisitions. In the 84 macular and optic-nerve cases, the layer segmentation chunks (TOP, BM, NFL [nerve fiber layer], GCL [ganglion cell layer], INL [inner nuclear layer]) were absent from the source .OPT file entirely: the device had not computed or stored on-device segmentation for those acquisitions, and no alternative chunk was available from which to derive the boundaries. This is not a signal-quality artefact: the scan quality index of the failing macular acquisitions is indistinguishable from that of the successful ones (median SQI 0.73 in both groups; range 0.32 to 0.85 versus 0.31 to 0.89), confirming that the shortfall reflects what the device wrote to disk rather than degraded acquisitions. Nor is it a parser limitation: Transducin extracts every quantitative finding the device actually stored. The remaining 8 were biometry acquisitions (of 10 at Site A) from which neither axial length nor central corneal thickness could be extracted; given the small number, this pathway was not pursued further. Site B, whose corpus contains a higher proportion of acquisitions with on-device segmentation present, reached 79/88 (89.8%). The Site A and Site B rates therefore differ by corpus composition and by what each device stored, not by version-dependent parser behaviour.

### 3.2 Measurement Validation

CMT (defined as mean ILM–BM distance within a 1 mm diameter circle, equivalent to ETDRS-C) was 283.9 µm for the OD scan (native SOCT central sector = 287 µm; difference = ™3.1 µm) and 291.8 µm for the OS scan (native SOCT central sector = 289 µm; difference = +2.8 µm). Both differences were within ±5 µm. The sub-±5 µm residual reflects the difference between Transducin’s pixel-grid circular integration and the SOCT device’s native interpolation method.

For axial length (from the MYOPI chunk), agreement was exact (to 2 decimal places in millimeters) for all 39 biometry files. For peripapillary RNFL, sector values agreed within ±1 µm for 92/94 cases, with the two discrepancies in low-SQI studies.

CMT was successfully extracted from 176 of 254 macular scans across four device/version groups. No statistically significant difference in CMT distribution was observed across SOCT versions or device models (Kruskal-Wallis H = 4.094, p = 0.2515; Figure 4), confirming parse-level format stability across software generations spanning approximately eight years of SOCT firmware evolution.

**Figure 4.**
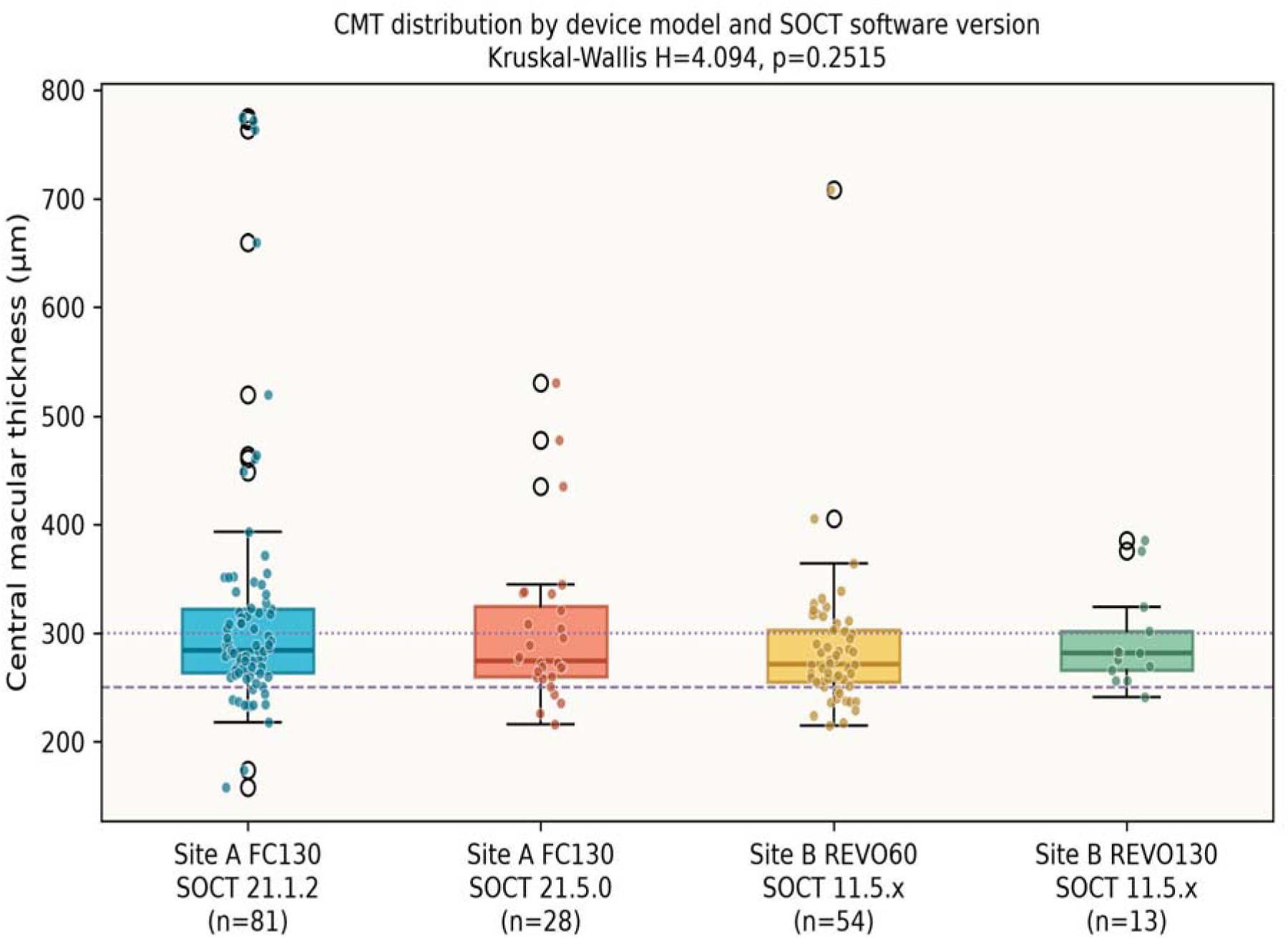
CMT distribution by device model and SOCT software version. Box plots show median (line), IQR (box), 1.5×IQR (whiskers), and individual data points (jittered). Reference lines at 250 μm (dashed) and 300 μm (dotted) indicate the normal foveal range. Four groups: Site A FC130 SOCT 21.1.2 (n=81), Site A FC130 SOCT 21.5.0 (n=28), Site B REVO 60 (SOCT 11.5.x, n=54), Site B REVO FC130 (SOCT 11.5.x, n=13). Kruskal-Wallis H = 4.094, p = 0.2515 (not significant), confirming format stability across software generations.

### 3.3 DICOM Conformance

All generated DICOM objects were validated using pydicom’s conformance checker with zero Type 1 attribute violations. All objects were successfully ingested by Orthanc 26.1.0 PACS [9] via C-STORE. OphthalmicTomographyImageStorage instances rendered correctly in OHIF Viewer v3 (Supplementary Video S1). SR instances were stored and retrievable via C-FIND but are not rendered visually by OHIF’s current ophthalmic viewer module — a known limitation of the viewer ecosystem, but could be opened as a text file or on other viewers such as Falcon MD (iCat Solutions, Norwich, UK).

### 3.4 Retroactive Processing

Retroactive SR generation for 11,956 historical series already stored in the clinical PACS was completed overnight using retroactive_sr.py, at approximately 800 studies per hour on the HP Z2 hardware with peak memory usage below 2 GB.

### 3.5 Round-trip Identity Validation

NOEL ID propagation was confirmed for all 387 files in the primary corpus. No identity mismatches were found between PatientID in generated DICOM objects and the corresponding EHR records. Figure 5 shows successful C-STORE delivery of all Transducin-generated DICOM objects.

**Figure 5.**
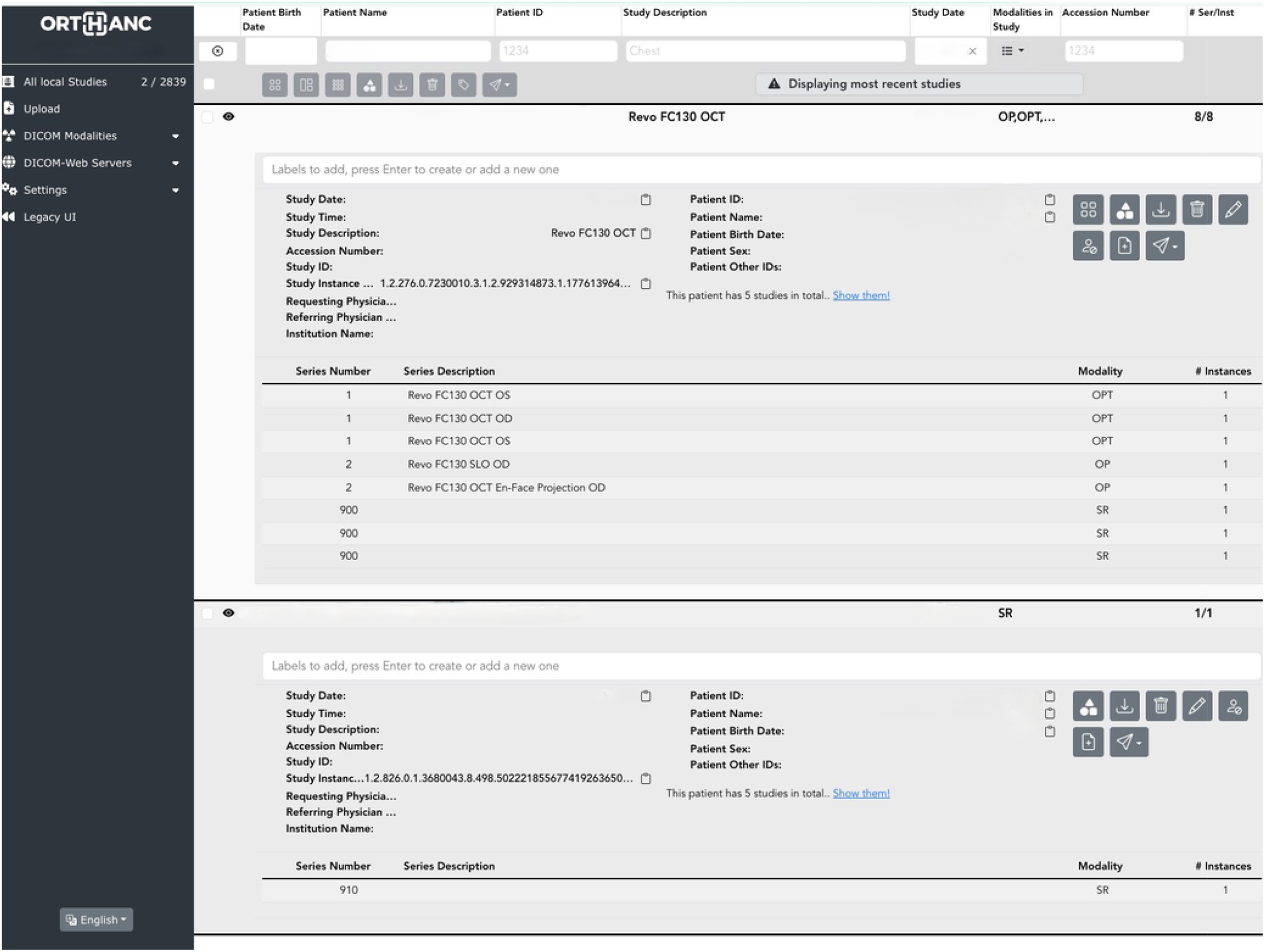
Orthanc PACS worklist, demonstrating successful C-STORE delivery of all Transducin-generated DICOM objects: OphthalmicTomographyImageStorage (OPT, ×3 series), OphthalmicPhotography (OP, ×2 — SLO and En-Face), and Structured Repor instances (SR, ×3). A separate SR-only study (Series 910) contains the macular TID 1500 Measurement Report with all SNOMED-CT coded findings.

### 3.6 Cross-Version and Cross-Model Validation

Transducin was applied to 88 additional .OPT files from Site B (REVO 60 and REVO FC130, SOCT v11.5.x), representing software versions approximately eight years prior to the primary corpus. Structural parsing succeeded for all 88 files (100%). Within a separate, curated 19-file pilot sub-study (13 REVO 60, SOCT 11.5.3, and 6 REVO FC130, SOCT 11.5.0; Table 4), one chunk-naming discrepancy was identified and resolved: SOCT 11.5.0 stores the Bruch’s membrane boundary under the key BOTTOM, whereas SOCT 21.1.2 uses BM. The parser was updated with a transparent fallback lookup:

**Table 4.** Site B cross-version validation summary (curated pilot sub-study: exact SOCT 11.5.0 vs 11.5.3 compariso, n=19).

| Device | SOCT version | Files (n) | Parse success | Macular studies (CMT extracted) | Laterality from tag 23 |
| --- | --- | --- | --- | --- | --- |
| REVO 60 | 11.5.3 | 13 | 13/13 (100%) | 8 (8/8) | 13/13 (100%) |
| REVO FC130 | 11.5.0 | 6 | 6/6 (100%) | 1 (1/1) | 5/5 applicable |
| Total | 11.5.0–11.5.3 | 19 | 19/19 (100%) | 9 (9/9) | 18/18 (100%) |

*The REVO FC130 macular scan (oct macular.opt) yielded a valid CMT; laterality was confirmable for 5 of the 6 FC130 files (the sixth being a biometry acquisition without a SLO pass).*

(_extract_layer_with_fallback).

Following this correction, within the 19-file pilot, CMT was successfully extracted from all 9 macular studies (8 REVO 60 and 1 REVO FC130).

Laterality inference from OCTPARAMS tag 23 was validated against all 13 REVO 60 files with ground-truth laterality labels embedded in filenames. The sign-based rule (positive → OS, negative → OD) achieved 13/13 accuracy (100%) with no ambiguous cases. The five REVO FC130 files without structured filenames were also assigned laterality from tag 23 and verified against the native SOCT display; all five assignments were confirmed correct. This demonstrates that the tag 23 laterality signal is present and consistent across both the REVO 60 and REVO FC130 hardware platforms and across both software versions tested (SOCT 11.5.0 and 11.5.3). Three files contained B-scan chunks written incompletely at export time; the parser skips these gracefully.

Beyond the curated 19-file pilot above, the recursive corpus discovery identified the complete Site B dataset: 88 .OPT files across two device models (REVO FC130 and REVO 60, SOCT v11.5.x), spanning all supported acquisition types (Table 4b).

**Table 4b.**
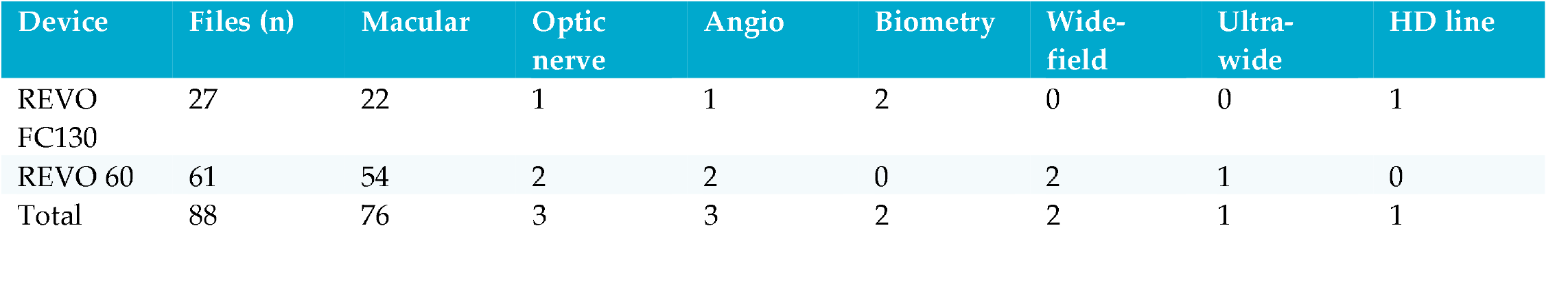
Site B — full discovered corpus (SOCT v11.5.x, n=88), by device and acquisition type.

| Device | Files (n) | Macular | Optic nerve | Angio | Biometry | Wide-field | Ultra-wide | HD line |
| --- | --- | --- | --- | --- | --- | --- | --- | --- |
| REVO FC130 | 27 | 22 | 1 | 1 | 2 | 0 | 0 | 1 |
| REVO 60 | 61 | 54 | 2 | 2 | 0 | 2 | 1 | 0 |
| Total | 88 | 76 | 3 | 3 | 2 | 2 | 1 | 1 |

### 3.7 Zeiss Cirrus HD-OCT Validation

The Cirrus pipeline was validated against 41 studies from three clinical datasets (Zeiss Cirrus HD-OCT 500). The corpus comprised 25 Macular Cube 512×128 studies and 16 Optic Disc Cube 200×200 studies (Table 5). SR generation succeeded for all 41 applicable studies (100%). The acquisition type was correctly identified via PerformedProtocolCodeSequence CodeValue in all cases. HD 5 Line Raster studies (n=56 in the full dataset) were correctly excluded from SR generation as they carry no analysis file.

**Table 5.** Zeiss Cirrus HD-OCT 500 validation corpus.

| Acquisition Type | Zeiss Code | Files (n) | SR Generated | Measurement range |
| --- | --- | --- | --- | --- |
| Macular Cube 512×128 | SD-MTA | 25 | 25 (100%) | CMT 203–630 $\mu\text{m}$ |
| Optic Disc Cube 200×200 | SD-GOUA | 16 | 16 (100%) | RNFL 53–123 $\mu\text{m}$; CDR 0.30–0.78 |
| Total |  | 41 | 41 (100%) |  |

CMT values ranged from 203 µm to 630 µm, with the upper extreme corresponding to clinically documented macular pathology. RNFL values of 53–63 µm in a subset of disc studies are consistent with glaucomatous damage; values of 95– 123 µm represent healthy or near-healthy nerve fiber layer. Cup-to-disc ratio (CDR) values of 0.30–0.78 span the clinically observed range. A dynamic layer dimension inference correctly handled both macular (128×512 pixels) and disc (200×200 pixels) ILM/BM arrays, resolving a previously undocumented format variation in the Zeiss SpatialRegistration file.

### 3.8 Forward Compatibility with SOCT 21.5.0

SOCT 21.5.0 (released April 2026) was verified against four .OPT files from a clinical REVO FC130 using two Ultra Wide Field (UWF) acquisition protocols acquired on the same device as the primary corpus. No new chunk types were introduced in 21.5.0 relative to 21.1.2, confirming the stability of the chunk-container architecture across this version range. UWF cube (256 × 768, ∼170 MB). Parse succeeded for both OD and OS. CMT (= ETDRS-C after circular-mask alignment) and ETDRS extraction were correct: OD 338.0 µm; OS 344.4 µm. pRNFL global (OD 29.7 µm, OS 29.8 µm), mGCIPL (OD 62.6 µm, OS 62.3 µm), and CDR (OD 0.567, OS 0.528) were extracted. Laterality was correctly inferred in all four files (100%). The DMARKERS chunk is absent in 21.5.0 UWF files, causing study_type misclassification as optic_nerve via the dimension-based fallback; scan_center_x (≈ ±24–45 mm vs. ±1–3 mm for standard macular acquisitions) provides a reliable secondary discriminator and is roadmapped for v1.1. UWF radial scan (8 × 2,560 A-scans, ∼27 MB). The radial star protocol stores 8 B-scans of 2,560 A-scans at ∼6.25 µm/pixel lateral resolution, covering ∼16 mm per line. The radial scan protocol does not include automated layer segmentation data; quantitative measurements (CMT, ETDRS) cannot be extracted. Because the file examined was only the .OPT, lacking ANALYSIS.DAT segmentation file, the radial scan protocol does not support SR file generation in v1.0.0 and is planned for future versions.

## 4. DISCUSSION

### 4.1 The Native DICOM Export Gap

Table 6 presents the gap between what the SOCT native DICOM export provides and what Transducin generates. The data in the left column are sourced entirely from the manufacturer’s own DICOM Conformance Statement v21.1.2 [1].

**Table 6.**
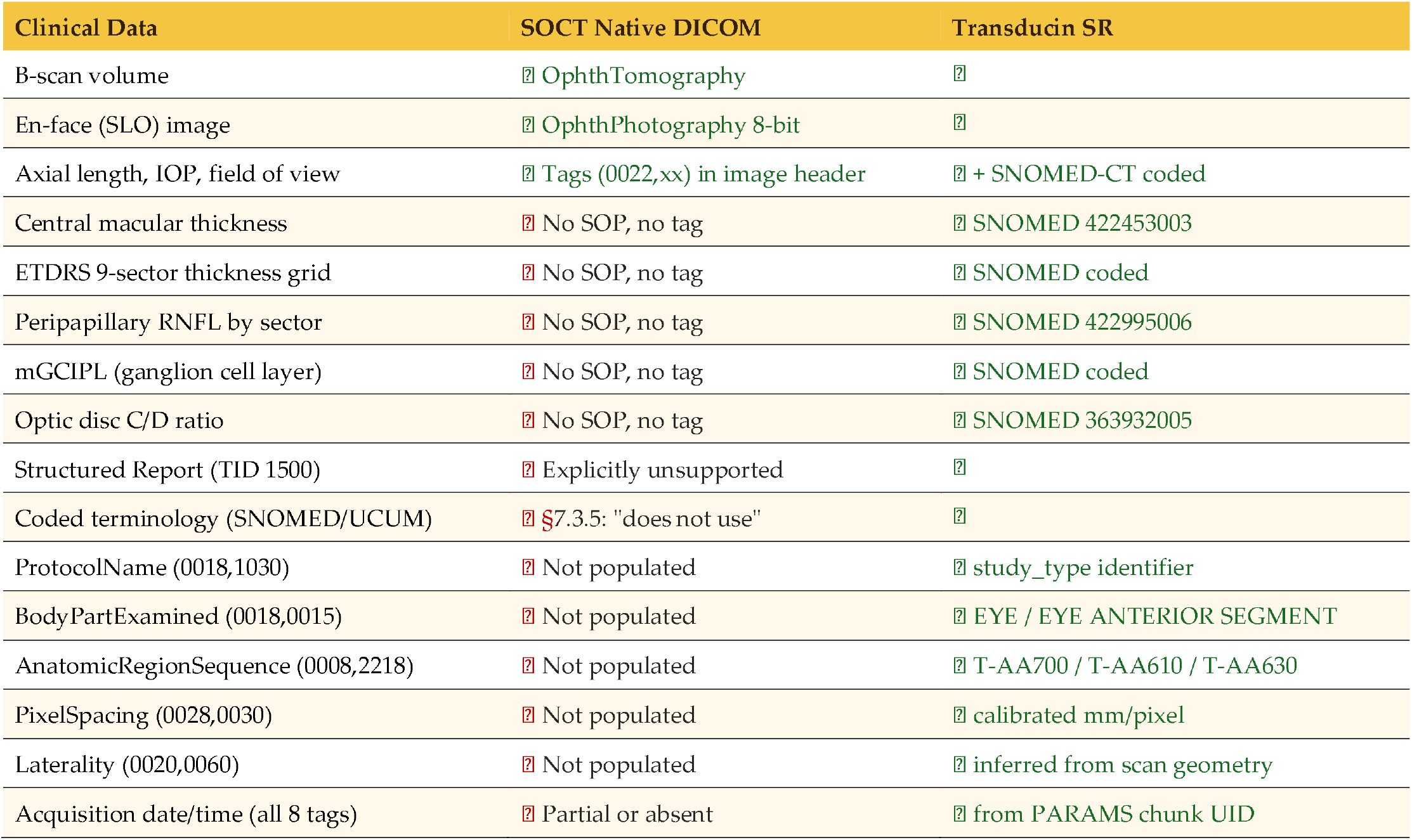
Native SOCT DICOM export versus Transducin SR output.

### 4.2 Laterality Inference: Geometric Encoding in Scan Acquisition

The discovery that OCTPARAMS tag 23 encodes laterality through the sign of the foveal horizontal position merits explicit discussion. The REVO FC130 computes this value from the SLO acquisition pass — specifically, the signed displacement in millimeters from the scan center to the foveal reflex or disc center detected during acquisition. Because the fovea is located nasal to the disc in both eyes, and because the nasal direction projects to opposite sides of the image in left versus right eyes, the sign of this displacement is a deterministic function of laterality that does not depend on any operator-entered metadata.

This has two practical implications. First, files exported without structured filenames (a common occurrence in older SOCT software versions or in workflows where the device operator does not follow a naming convention) can be reliably assigned laterality without human intervention. Second, the same principle likely applies to other OCT devices that store a foveal or disc center position in their acquisition metadata, suggesting that geometry-based laterality inference may be more broadly applicable across the ophthalmic imaging device landscape than previously recognized. Whether Heidelberg, Zeiss, or Topcon devices encode an analogous parameter in their proprietary formats is beyond the scope of this work but represents a natural extension.

### 4.3 Scan Type Discrimination via Chunk Architecture

The identification of DMARKERS as a device-invariant discriminator between optic nerve and macular acquisitions demonstrates a general principle: the chunk architecture of the .OPT format encodes acquisition semantics that are more reliable than any external metadata. The DMARKERS chunk is present exclusively in disc-centered acquisitions because disc coordinate detection is an integral step in the REVO FC130’s disc analysis pipeline — it exists only when the acquisition firmware has detected and localized the optic disc. This makes its presence a ground-truth signal for disc acquisition type, invariant to software version, device model, or operator workflow.

The complete scan type inference hierarchy (Section 2.3) follows a similar principle: each level of the hierarchy uses the most reliable available signal, falling through to less reliable signals only when necessary. This design produced zero classification errors across 475 files from four software versions and three device models.

### 4.4 Comparison with Related Work

OCT-Converter [2] provides Python parsing for Heidelberg (.e2e, .vol), Zeiss (.img, .ex.dcm), Topcon (.fda, .fds), Optovue, and Bioptigen formats, but explicitly does not support any Optopol format. eyepy [3] focuses on Heidelberg .vol files with layer segmentation. Neither library has ever included Optopol support. The present work fills this gap.

Regarding DICOM SR for ophthalmic OCT, the highdicom library [7] provides TID 1500 support but no ophthalmic-specific templates and no Cirrus or Optopol extraction logic. The present work contributes the first real-world validated use of TID 1500 for quantitative ophthalmic measurements across two major OCT platforms.

Regarding the Cirrus platform specifically, no prior open-source tool converts Cirrus private-tag measurements to TID 1500 SRs with SNOMED-CT coded findings. The 99CZM coding scheme used by Zeiss is not publicly documented; its structure was characterized empirically in this work.

The 2024 launch of AI-READI (the first publicly available, standards-compliant DICOM retinal imaging dataset, developed under the NIH Bridge2AI program) demonstrates growing institutional momentum toward the same interoperability goal pursued here at a much smaller scale [4,10]. Three gaps nonetheless remain even in that flagship effort. First, device coverage: AI-READI’s OCT arm spans Heidelberg Spectralis, Topcon Maestro2/Triton, and Zeiss Cirrus, but not the Optopol REVO platform addressed by Transducin. Second, methodological transparency: AI-READI’s public documentation states only that exported files were “formatted according to the NEMA DICOM standards,” without disclosing the source proprietary formats or the conversion tooling itself — the standardization pipeline remains a closed, institutional process. Third, scope: AI-READI standardizes pixel data and acquisition metadata but does not extend to structured clinical reporting. Transducin’s .OPT parser and Cirrus private-tag interpreter are released as open-source software with the reverse-engineering methodology documented as the primary contribution, and extend standardization through to TID 1500 Structured Reports carrying the derived quantitative measurements themselves rather than image data alone. Open, device-specific pipelines of this kind are complementary to consortium-scale efforts such as AI-READI: the latter provides breadth and institutional backing, while independently published, clean-room parsers can extend coverage to devices and vendors that any single consortium has not yet reached, without waiting for manufacturer cooperation. We reviewed AI-READI’s public documentation and dataset structure but did not attempt to independently identify or reverse-engineer its underlying acquisition metadata schema, as the consortium’s source files and conversion tooling were not accessible to us.

### 4.5 Limitations

- **Structured Report generation rate**. The gap between the Site A (51.2%) and Site B (89.8%) SR generation rates reflects differences in on-device segmentation availability across the two corpora, not a parser limitation; the full quantitative breakdown is reported in Section 3.1. Recovering the segmentation-absent files would require Transducin to perform its own retinal layer segmentation, which falls outside the scope of a format-translation tool and is left to future work.
- **Cross-site validation scope**. The primary corpus was acquired at a single site (387 .OPT files, SOCT v21.1.2 and v21.5.0). Cross-version compatibility was confirmed against 88 .OPT files from two devices at Site B. Validation against other SOCT device family models (REVO HR, NX 130, REVO 80) and versions between 11.5.x and 21.1.x was not performed.
- **SOCT 21.5.0 forward compatibility**. Four UWF .OPT files from SOCT 21.5.0 were processed. UWF cube acquisitions (256 × 768): parse succeeded with correct CMT/ETDRS extraction and laterality inference (100%). DMARKERS chunk absent in 21.5.0 UWF files causes study_type misclassification via dimension fallback; fix roadmapped for v1.1. UWF radial scan (8 × 2,560): no ANALYSIS.DAT segmentation present (intentionally excluded in this study); SR generation not supported in v1.0.0. No new chunk types introduced in 21.5.0 vs. 21.1.2.
- **Bruch’s membrane chunk naming**. SOCT 11.5.0 uses BOTTOM; SOCT 21.1.2 uses BM. Resolved via transparent fallback. Additional version-specific naming variations cannot be excluded.
- **Laterality from tag 23 in zero-value cases**. OCTPARAMS tag 23 carries a value of 0.0 mm for biometry acquisitions which lack a SLO pass. These files correctly receive no laterality assignment.
- **Biometry chunk schema variation**. SOCT <21.x uses WTW/TPR/S0–S9 chunks rather than MYOPI JSON. Biometric extraction not supported for pre-21.x files.
- **BOTTOM layer segmentation errors in chorioretinal pathology**. In two Site-B studies, systematic CMT overestimation (Δ >76 µm) was caused by the SOCT segmentation algorithm anchoring the BOTTOM boundary to pathological structures rather than Bruch’s membrane. In SITE_B_010 (dry AMD with large drusen), BOTTOM tracked the outer limit of the RPE–drusen complex rather than true Bruch’s membrane; the flat dip profile (dip=17 µm) is a reliable diagnostic indicator of this failure mode. In SITE_B_013 (sub-RPE fibrosis after photocoagulation), BOTTOM traced the fibrotic layer across the central fovea, producing a bimodal thickness distribution (250–275 µm pericentral vs. 300–350 µm in the photocoagulated zone). Transducin faithfully reproduces the SOCT segmentation output and therefore inherits these errors. Downstream users should flag scans with dip <20 µm for manual BOTTOM layer review before including CMT values in quantitative analyses.
- **Truncated B-scan frames**. Three Site B files contained B-scan chunks written incompletely at export time, skipped gracefully.
- **Cirrus HD 5 Line Raster SR exclusion**. Cirrus HD 5 Line Raster SD-S51 acquisitions generate DICOM image objects but no analysis file and therefore no SR output. This is a Cirrus platform constraint, not a Transducin limitation.
- **Cirrus acquisition type encoding**. The CodeValue/CodeMeaning in PerformedProtocolCodeSequence uses the private Zeiss coding scheme 99CZM, which is not publicly documented. The mapping characterized in this work (SD-MTA, SD-GOUA) covers the two acquisition types with analysis data; additional Cirrus acquisition types may require characterization.
- **Anterior segment OCT classification**. When filenames follow the standard REVO export convention without protocol keywords, anterior and posterior wide-field acquisitions cannot be automatically distinguished at the .OPT level — no chunk discriminator for this case exists in the versions tested. Classification defaults to posterior; users acquiring anterior segment OCT should include a recognizable keyword (e.g., “topo”, “anterior”) in the filename.
- **B-scan display fidelity**. Formal quantitative validation of pixel-level fidelity between extracted B-scans and SOCT native display is beyond the scope of this work.
- **OHIF SR rendering**. Current OHIF Viewer does not render TID 1500 ophthalmic measurement values visually, but they are generated and sent to the PACS on the transducing pipeline ready for querying. The SR files could successfully be opened graphically using Falcon MD Software.
- **Maintenance and version drift**. Reverse-engineered parsers are inherently vulnerable to silent breakage when a vendor revises its firmware or export format without notice. Transducin mitigates this risk in three ways. First, the fixture-based regression suite (tests/fixtures/) is run against every commit and pull request via continuous integration, so a firmware-driven change that alters chunk structure or byte layout is expected to surface as a test failure rather than a silent misparse. Second, the cross-version validation reported in Sections 3.1 and 3.6 (SOCT 11.5.0 through 21.5.0, spanning approximately eight years) demonstrates that the underlying chunk-container architecture has in practice remained stable across multiple vendor software revisions; new SOCT releases are added to this validation corpus as they become available in the authors’ own clinical practice, which continues to generate .OPT files on a daily basis and thereby provides an ongoing, real-world drift signal. Third, Transducin is released under an open-source license (Apache 2.0) with public issue tracking, so that any user encountering a format change in an untested SOCT or Cirrus firmware version can report it and, where the chunk-based architecture documented here still applies, contribute a fix without depending on the original authors’ continued access to that specific hardware and software combination. Collaboration is the basis for software development.

### 4.6 Forward Compatibility with DICOM Supplement 247 and Broader Standards Landscape

DICOM Supplement 247 (Eyecare Measurement Templates) was incorporated into the DICOM standard in the 2025b release (July 2025) [11]. It defines TIDs 2100–2124 specifically for ophthalmic structured reporting, superseding the older TID 4230/4231/4241 family. The current implementation uses SNOMED-CT codes within TID 1500, which imposes no restriction on coding scheme. Migration to TID 2123 and TID 2124 will require adopting the LOINC codes defined in CID 4285 for macular thickness measurements and the DCM codes defined in CID 4283 for circumpapillary RNFL measurements, as specified in Supplement 247. The provisional 99OFTALMOS codes in the current release are tagged for replacement with these official codes upon completion of the planned highdicom pull request. The container template for these structured reports is TID 2120 (Ophthalmic Measurements), with finalized DCM codes 131242/131243 replacing the draft-stage provisional identifiers; individual RNFL findings now carry final DCM codes 131264–131272 and 131274 (circumpapillary ROI width) or 131276–131287 (12 clockface positions), and macular findings carry DCM code 131255. Laterality coding also moved from the draft CID 244 to the ratified CID 247. As of this writing, highdicom’s own implementation of these ratified templates (tracked in pull request #407, the same pull request referenced in Section 2.6) remains open and unmerged; Transducin’s provisional 99OFTALMOS scheme therefore remains in active use pending that library update.

Supplement 247, while a significant advance, does not currently define measurement templates for several modalities present in the Transducin validation corpus. Ultra-wide field OCT has no dedicated peripheral retinal measurement templates; such acquisitions fall under the existing OphthalmicTomographyImageStorage SOP class (1.2.840.10008.5.1.4.1.1.77.1.5.4, Supplement 110 [12]) and are distinguished from standard-field acquisitions via the Horizontal Field of View tag (0022,000C). OCTA data are covered by dedicated SOP classes in Supplement 197 [13] (Ophthalmic OCT B-scan Volume Analysis Storage, 1.2.840.10008.5.1.4.1.1.77.1.5.8; Ophthalmic OCT En Face Image Storage, 1.2.840.10008.5.1.4.1.1.77.1.5.7), but Supplement 247 does not include corresponding measurement templates for vascular density, foveal avascular zone area, or perfusion index. Biometric OCT measurements are addressed by the Ophthalmic Axial Measurements Storage Image Object Definition (IOD) 1.2.840.10008.5.1.4.1.1.78.7 defined in Supplement 144 [14], which is not referenced by Supplement 247. Corneal topography was originally planned as TID 6007 within Supplement 247 but was removed from the final release and deferred to a future revision [11]; the existing Corneal Topography Map Storage SOP class (1.2.840.10008.5.1.4.1.1.82.1, Supplement 168 [15]) provides image storage without a companion SR template.

These gaps are consistent with an asymmetric maturity in the DICOM ophthalmic standard ecosystem, in which image storage IODs substantially precede their corresponding structured reporting templates. Transducin’s multimodal clinical corpus (spanning macular, disc, OCTA, wide-field, ultra-wide-field, biometry, and anterior segment acquisitions) provides real-world validation data that may support the development of missing templates through engagement with DICOM Working Group 09, highdicom and the HL7 FHIR Eyecare Implementation Guide [16]. This design choice anticipates DICOM’s own trajectory for AI-generated content: Supplement 219, developed by Working Group 23 (Artificial Intelligence/Application Hosting), defines a JSON representation for structured reports built directly on the TID 1500 Measurement Report template used here, underscoring TID 1500’s position as the template of choice for AI-derived ophthalmic measurements going forward [4].

### 4.7 Clinical and Research Implications

Transducin enables practices operating REVO 60, REVO FC130, or Cirrus HD-OCT 500 to integrate their quantitative OCT data into any DICOM-compliant PACS with full machine-readable measurements available for query, retrieval, and display. This is of particular relevance to all practices, but more so in resource-constrained settings where commercial integration solutions are economically inaccessible. A related benefit is the reduction of manual data-entry burden. Large-scale real-world retinal disease registries such as Fight Retinal Blindness! (FRB!) address this by minimizing, rather than eliminating, manual entry: a single structured encounter, five minutes for a new patient and two minutes for follow-up, in which a clinician or coordinator still transcribes values such as CMT into the registry by hand [17]. Transducin addresses the same underlying problem one step further upstream: because CMT, ETDRS, RNFL, and other quantitative findings are extracted directly from the device’s own raw output and encoded as machine-readable SR content, no manual transcription step is required at all — the measurement flows from acquisition to PACS-queryable, SNOMED-coded structured report without a human intermediary at any point.

The multivendor architecture generates identical TID 1500 SRs with SNOMED-CT coded findings regardless of whether the source device is an Optopol REVO or a Zeiss Cirrus (and quite possibly others). This enables longitudinal research studies that span device transitions, a common occurrence in clinical practice. The same PACS query retrieving CMT values by SNOMED code 422453003 returns measurements from both devices without vendor-specific filtering as needed (Figure 6). This multivendor scope is expanding: as described in Section 2.8, Transducin already generates TID 1500 visual field Structured Reports from Optopol PTS 925Wi static perimetry studies via a dedicated PACS watcher, extending coded interoperability beyond OCT to a second acquisition modality. Support for additional proprietary imaging formats, including Heidelberg Spectralis (.e2e), Topcon (.fda/.fds), and Bioptigen (.OCT), is similarly planned via the oct-converter library [2] as an image-extraction front end feeding the same TID 1500 SR pipeline and plans are on the pipeline for other ophthalmic imaging modalities.

**Figure 6.**
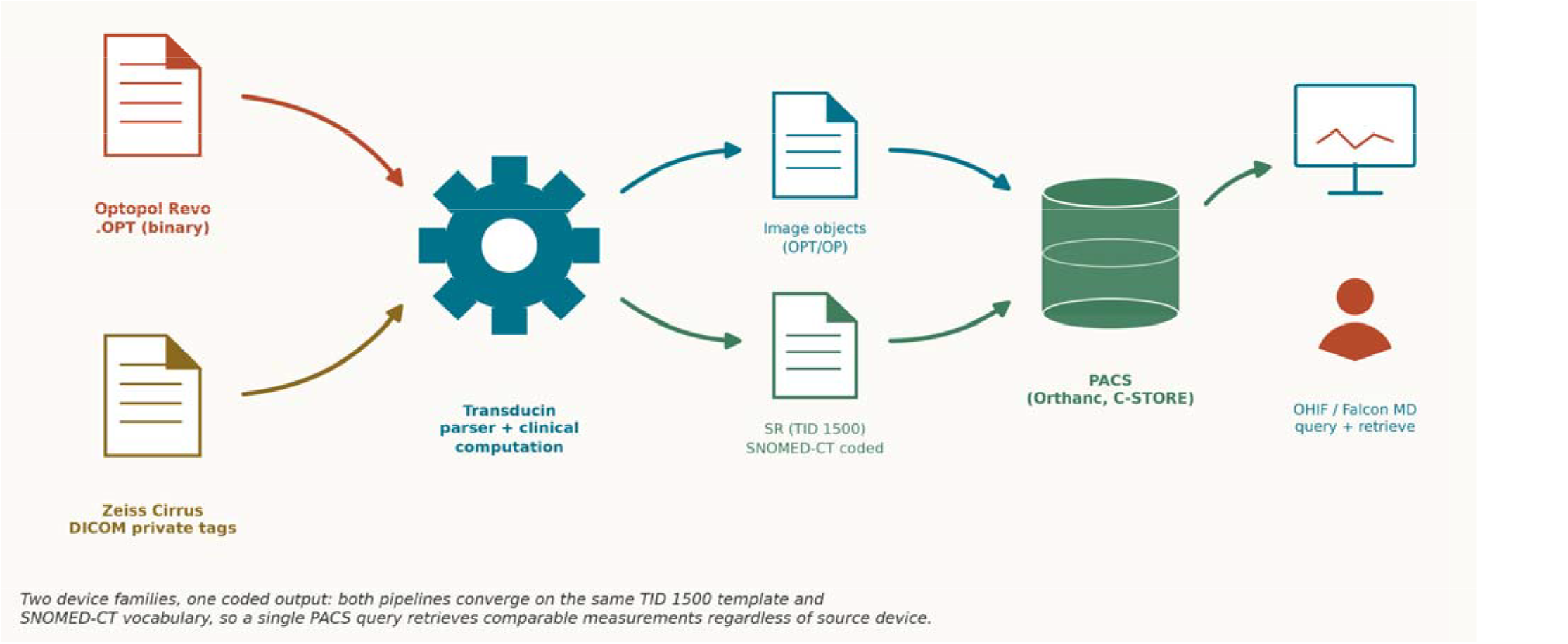
Transducin multivendor pipeline, from acquisition file to PACS query. Two device families (Optopol REVO .OPT and Zeiss Cirrus DICOM private tags) are parsed by the same Transducin pipeline into DICOM image objects and a SNOMED-CT coded TID 1500 SR, stored in Orthanc via C-STORE and retrievable by any standards-compliant viewer (OHIF, Falcon MD) without vendor-specific tooling.

The confirmation of format stability across SOCT versions 11.5.0 through 21.1.2 enables retroactive application of Transducin to historical archives accumulated under older software versions. The 11,956-study corpus now available in the clinical PACS at Site A represents an immediately actionable research dataset. The NOEL ID protocol demonstrates a generalizable approach to patient identity federation across heterogeneous clinical systems in settings where national health identifiers are unavailable or impractical.

### 4.8 Legal and Ethical Considerations Regarding Interoperability Analysis

The devices examined in this study were lawfully acquired and operated as part of routine clinical practice at the authors’ institutions. Neither author was presented with or accepted an end-user license agreement (EULA), non-disclosure agreement, confidentiality obligation, or other contractual restriction specifically governing analysis of the relevant file formats. Format characterization was performed exclusively on data generated by the authors’ own institutions and with reference to publicly available or manufacturer-supplied documentation, including the Conformance Statement [1] and CMDL Interface documentation [6]. No non-public vendor documentation, proprietary source code, credentials, or confidential vendor communications were obtained or used.

The limited single-byte XOR sweep described in Section 2.1 was performed solely to determine whether the unidentified .OPT patient-identity block used trivial single-byte obfuscation. That block contains factual patient-identification data rather than original expressive content, which raises a threshold question as to whether copyright-based technological-protection-measure frameworks apply to it at all, independent of the outcome of the test described below. The test produced no intelligible patient information, no data or key were recovered from the block, and no further cryptanalytic or circumvention procedure was attempted. Because the test neither decrypted the block nor otherwise avoided, bypassed, removed, deactivated, or impaired the mechanism protecting it, that protection remained effective and the block remained inaccessible throughout the analysis; patient identity is instead established independently through the NOEL ID protocol described in Section 2.7. The Zeiss Cirrus pixel-data operation examined in Section 2.10 appeared to constitute a reversible byte-level transformation rather than an authentication or access-control mechanism. The transformation had previously been described in a public technical discussion [8], and its implementation in Transducin does not incorporate proprietary vendor source code.

The .OPT parser was developed through independent analysis of lawfully generated binary files together with the publicly available manufacturer documentation cited above. No confidential format specification or non-public implementation information was used. Independent precedent for technical analysis of proprietary OCT file formats has also been reported in the scientific literature [18]. This work was conducted in Mexico. Article 164 of Mexico’s Federal Law for the Protection of Industrial Property (Ley Federal de Protección a la Propiedad Industrial, LFPPI) provides that independent discovery or creation, and the observation, study, disassembly, or testing of a product that has been made publicly available or is lawfully possessed by the person obtaining the information, do not constitute trade-secret misappropriation when no applicable confidentiality obligation exists. Article 114 Quáter of the Federal Copyright Law (Ley Federal del Derecho de Autor, LFDA) likewise recognizes specified good-faith, non-infringing reverse-engineering activities involving lawfully obtained computer software when undertaken for the sole purpose of achieving interoperability with independently created software. These provisions provide relevant legal context for the methodology described here but are not presented as a formal legal opinion or as a definitive determination of the legal status of any particular operation. Laws governing reverse engineering, technological protection measures, contractual restrictions, patents, and trade secrets vary among jurisdictions, and persons undertaking similar work should evaluate the law and contractual terms applicable in their own setting. Institutional ethics approval covered the clinical-data validation component of this work; it did not constitute legal review of the interoperability-analysis methodology. A preprint of the study was made publicly available on medRxiv on 14 July 2026 and was subsequently provided to Optopol Technology Sp. z o.o. Transducin is distributed under the Apache License, Version 2.0. Section 3 of that license provides users with a patent license from each contributor for patent claims licensable by that contributor that are necessarily infringed by that contributor’s contribution, subject to the terms of the license. No patent application has been filed or sought by the authors covering Transducin or the methods reported here. The Apache 2.0 patent license applies only to qualifying patent claims licensable by contributors and does not extend to patents held by unrelated third parties.

The purpose of Transducin is interoperability: it enables clinical imaging data generated by lawfully operated instruments to be represented as standardized DICOM objects for archival, analysis, and research use. It does not reproduce vendor application software, provide access to vendor source code, recover the inaccessible patient-identity block described above, or bypass device authentication. Transducin should therefore be understood as an independently developed interoperability tool rather than as a replacement for, modification of, or means of obtaining unauthorized access to the manufacturers’ software systems.

Two recent, unrelated developments illustrate potential future directions rather than current dependencies of Transducin. Multimodal foundation models for retinal OCT analysis, such as MIRAGE [19], currently operate on unstructured pixel data; the standardized, coded measurements generated by Transducin’s SR pipeline could in principle support future calibration of such embedding-based models against ground-truth clinical quantities. A related example is provided by population-scale retinal embeddings (Ret-AAE) trained on UK Biobank OCT and fundus photographs, recently inked to cardiovascular and neurodegenerative disease risk [20], and by fundus-photograph screening models for conditions such as ADHD in children [21]; in both cases the predictive features are learned rather than clinically coded, and coded quantitative retinal output of the kind Transducin generates could serve as an interpretable comparison arm. These oculomics-style applications typically draw on multiple imaging modalities; the REVO FC130 platform is itself broadly multimodal but does not include fundus autofluorescence (FAF), a modality present in cohorts such as AI-READI [10], so the corpus accessible via Transducin does not fully replicate the modality panel used by some oculomics studies. Separately, Optical character recognition (OCR) pipelines for legacy OCT reports, such as OCTess [22], point toward a complementary future extension of this broader software ecosystem for ingesting historical non-DICOM archives; Transducin does not currently implement OCR-based extraction.

Beyond immediate interoperability, Transducin’s contribution aligns with a broader call from academia to treat data-liberation infrastructure (not only novel model architectures) as a legitimate form of scholarship [23]. Proprietary OCT formats are a documented barrier to model retraining and lifecycle management, forcing researchers at under-resourced institutions to either reverse-engineer vendor formats independently or forgo retrospective data entirely. By converting Optopol .OPT and Zeiss Cirrus private-tag data into standard DICOM SR, Transducin lowers this barrier for smaller practices, such as the two-site collaboration described here, to participate in the kind of diverse, cross-institutional dataset-building that mitigates spectrum and demographic bias in medical AI. The cross-version stability demonstrated in Section 3.6 across four SOCT software versions spanning approximately eight years directly addresses the software-drift concern raised as a barrier to retraining, transforming previously siloed archives into standardized, reusable research assets.

As tools like Transducin lower barriers to data liberation and sharing, the associated risks of model inversion and membership inference attacks warrant explicit consideration [23]. These risks, however, pertain specifically to trained machine learning models capable of memorizing patterns from their training data — Transducin itself performs no learning; its outputs are produced deterministically from documented binary structure and geometric rules (e.g., the sign-based laterality inference described in Section 3.5), with no fitted parameters and therefore no mechanism by which an individual patient’s data could be reconstructed or their membership inferred from the software itself. This distinction matters as Transducin-generated SR becomes an input to downstream AI development: models trained on Transducin-extracted measurements should adopt privacy-preserving training strategies (e.g., differential privacy), avoid publishing per-patient embeddings or full model weights without safeguards, and apply minimum cell-size thresholds when reporting cohort demographics.

## 5. CONCLUSION

Transducin provides the first publicly documented specification of the Optopol REVO FC130 .OPT binary format and is the first open-source multivendor pipeline generating TID 1500 DICOM Structured Reports with SNOMED-CT coded ophthalmic measurements from both Optopol REVO 60 and FC130, as well as the Zeiss Cirrus HD-OCT 500 devices. For the Optopol platform, the contribution is doubly confirmed by the absence of any prior publication on this format and by the manufacturer’s own Conformance Statement acknowledging the absence of coded terminology. For the Cirrus platform, the contribution is the first characterization of the private 99CZM coding scheme and extraction of quantitative measurements into standards-compliant SRs.

Beyond the format parsing contribution, this work demonstrates that OCT devices embed geometric signals, specifically the signed horizontal position of the foveal or disc reflex, that reliably encode laterality independent of operator data entry. This finding is validated with 100% accuracy across 18 files from two device models and two software versions, and suggests a broadly applicable principle for automated laterality inference in ophthalmic data pipelines. All of these, we hope, will be further validated or expanded upon by the scientific and software communities. Cross-version validation across SOCT versions 11.5.0 through 21.1.2 demonstrated format stability spanning approximately eight years, with the DMARKERS chunk serving as a device-invariant discriminator for optic nerve versus macular acquisitions across all tested configurations. All code is available at https://github.com/oftalmos-org/transducin under the Apache License 2.0.

## Supporting information

Supplementary video 1. Transducin_in_OHIF_example_video

## Data Availability

The source code is available at https://github.com/oftalmos-org/transducin under the Apache License 2.0. De-identified cross-site validation results are available at validation/site_b_results_anon.csv in the public repository. De-identified synthetic .OPT files for format validation are available at tests/fixtures/ in the repository. Primary patient data cannot be shared due to Mexican NOM-004-SSA3-2012 privacy regulations.

https://zenodo.org/records/20031717

## Supplementary Video S1 (OHIF Viewer rendering of Transducin-generated DICOM image series) is available in the repository at docs/demo/Transducin_in_OHIF_example_video.mp4

## Ethics statement

The retrospective validation corpus was drawn from routine clinical data under Mexican NOM-004-SSA3-2012 institutional data use provisions. This work is a software development and validation study. No patient data are distributed. An institutional ethics committee (Hospital Angeles Chihuahua) waived ethical approval for this work.

## AI Tools Disclosure

Portions of the research for Transducin, its codebase, data validation scripts, and manuscript text were developed with the assistance of large language model tools (LLMs), including Claude (Anthropic), ChatGPT (OpenAI), Google Antigravity/Gemini (Google), and locally hosted LLMs via LM Studio or Ollama. These tools were used for code development and debugging, data validation script generation, and editorial assistance in manuscript preparation. All AI-assisted outputs were reviewed, verified against source data and code, and edited by the author, who takes full responsibility for the accuracy, integrity, and originality of the final manuscript and software.

## Declarations

### Ethics Approval and Consent to Participate

The retrospective validation corpus was drawn from routine clinical data obtained during standard-of-care visits under Mexican NOM-004-SSA3-2012 institutional data use provisions. This work is a software development and validation study. No patient data are distributed. An institutional ethics committee (Hospital Angeles Chihuahua) waived ethical approval for this work.

### Consent for Publication

All validation data are fully de-identified. The sole identifiable dataset (macular OCT acquisition belonging to the corresponding author) was obtained with explicit personal informed consent for use and publication as a validation case.

### Availability of Data and Materials

Source code is publicly available at https://github.com/oftalmos-org/transducin under the Apache License 2.0. The de-identified cross-site validation dataset is archived on Zenodo (doi:10.5281/zenodo.20031717). Patient data cannot be shared due to Mexican NOM-004-SSA3-2012 privacy regulations. Synthetic .OPT fixture files for format validation are available in the repository at tests/fixtures/.

### Authors’ Contributions

Jaurrieta Hinojos JN (corresponding): Conceptualization, Methodology, Software, Validation, Formal analysis, Investigation, Data curation, Writing – original draft, Writing – review & editing, Visualization, Supervision, Project administration. Palomares Ordonez JL: Investigation (Site B data acquisition), Clinical validation, Writing – review & editing. Chacon Hinojos JF: Software (Zeiss Cirrus decoder), Validation, Formal analysis, Writing – review & editing. Folgueras Batres MA: Software (PACS integration, deployment infrastructure), Writing – review & editing.

## Acknowledgements

The author thanks the staff of Oftalmos, the Retina and Vitreous practice at Hospital Angeles Chihuahua, for their support throughout this work. Special thanks are due to L. Robles, MBA, for collaboration on legal and strategic matters relating to the open-source release and licensing framework; to L. Zertuche, for assistance with image acquisition; and to C. Solis, for administrative coordination at the practice. The author also thanks the Research Ethics Committee of Hospital Angeles Chihuahua for its review of the study protocol. The author acknowledges the open-source software on which Transducin depends: pydicom and pynetdicom (D. Mason and contributors) for DICOM manipulation and network operations, highdicom (M.D. Herrmann and contributors) for Structured Report generation, and the Orthanc PACS ecosystem (S. Jodogne). The design of Transducin was inspired by OCT-Converter (M.S. Graham and contributors), which pioneered open-source parsing of proprietary OCT formats, and by OCTess (M. Balas), which informed the approach to extracting structured clinical data from Cirrus PDF reports. The author is grateful to C.P. Bridge for guidance on structuring Transducin’s output within the highdicom TID 1500 framework. The author also thanks DICOM Working Group 09 (Ophthalmology) for crafting Supplement 247 and David Hockney for the colour palette inspiration for this manuscript. The authors also thank Dr. David A. Clunie (PixelMed Publishing) for reviewing the preprint and identifying errors in references [2] and [3], and more broadly for his foundational contributions to the DICOM standard on which this work depends.

